# Healthcare-resource-adjusted vulnerabilities towards the 2019-nCoV epidemic across China

**DOI:** 10.1101/2020.02.11.20022111

**Authors:** Hanchu Zhou, Jiannan Yang, Kaicheng Tang, Qingpeng Zhang, Zhidong Cao, Dirk Pfeiffer, Daniel Dajun Zeng

**Author notes:** Correspondence to (Qingpeng Zhang), (Daniel Dajun Zeng). These first authors contributed equally to the study (Hanchu Zhou, Jiannan Yang, Kaicheng Tang).

## Abstract

We integrate the human movement and healthcare resource data to identify cities with high vulnerability towards the 2019-nCoV epidemic with respect to available health resources. The results inform public health responses in multiple ways.

## Introduction

The 2019-nCoV epidemic has spread across China and 24 other countries^1–3^ as of February 8, 2020. The mass quarantine measure implemented in the epidemic’s epicenter, Wuhan, Hubei Province, China, on January 23, 2020, has stopped the human migration between Wuhan and other places. However, according to the local authorities, over five million people had already traveled from Wuhan to other parts of China in the weeks leading to January 23, 2020, with the majority (79.96%) to the other cities within Hubei.

Healthcare resources have become a major bottleneck in the response to this public health crisis. The sudden onset of the epidemic and the sheer size of the infection have caused significant shortage in healthcare resources across Hubei Province. So far, there have been over 11,000 non-local healthcare professionals, mobilized by the public health authorities from other parts of China, fighting against the epidemic in Wuhan. However, this is far from adequate for Wuhan given the severity of the epidemic. In addition to Wuhan, other cities in Hubei are in need of urgent external help as well with respect to health resources, as multiple localized outbreaks are taking place. On February 7, 2020, the National Health Commission of China (NHCC) initiated the “One City Covered by One Province” program, explicitly assigning a specific province to aid one city in Hubei. Two critical questions of operational nature remain, which cities, both in Hubei and other provinces, are most vulnerable to localized outbreaks, and how the public health authorities should balance the healthcare resources under stress to best fight the epidemic and minimize the risks? This study is aimed to shed light on these questions by developing a vulnerability analysis framework integrating the human movement and healthcare resource data.

## Methods

This study makes use of four public datasets: (a) the daily counts of confirmed 2019-nCoV cases, collected from NHCC^4^; (b) the list of designated hospitals with the number of reserved beds, retrieved from the websites of the provincial and municipal health commissions; (c) the information of the hospitals designated for 2019-nCoV, retrieved from 99.com.cn, a public hospital database; and (d) the human movement data, provided by the Baidu Migration (http://qianxi.baidu.com/), a public site offering real-time migration information across mainland China.

Recent studies indicated that the confirmed case data for Hubei Province, in particular, Wuhan, was subject to the under-reporting bias, while the data from places outside of Hubei is more reliable for inferring the actual epidemic size^5^. Following Wu *et al*.^5^, we developed a susceptible-exposed-infectious-recovered (SEIR)-based metapopulation model, which has 100 sub-populations representing the 100 cities that have the greatest number of people traveling from Wuhan. Note that we adopted the same epidemiological parameters as Wu *et al*.^5^. This model was then used to estimate *ID*_*i*_, the number of infected individuals in each city. Lastly, healthcare-resource-adjusted vulnerability indexes to the 2019-nCoV outbreak for city *i, V*_*i*_ were obtained as follows: *V*_*i*_ *= ID*_*i*_ */ B*_*i*_, where *B*_*i*_ represents the total number of reserved beds in the designated hospitals.

## Results

The Figure presents the estimated number of infected individuals, as well as the healthcare-resource-adjusted vulnerability to the 2019-nCoV outbreak across mainland China, as of January 29, 2020. The epidemiological parameters may have changed after January 23, 2020, when the lockdown measure was implemented. Thus, we only use the data by January 23, 2020 and produce the estimation till January 29, 2020 to accommodate the six-day incubation period identified by Wu *et al*.^5^.

The ten most vulnerable non-Hubei cities are also listed in the figure. Note that the cities in Hubei (which are being aided by other provinces) are not presented here because of their significantly higher vulnerability as compared to others. Among the ten most vulnerable non-Hubei cities, Changsha, Beijing, Chongqing and Shanghai are major metropolises with a large number of estimated infected individuals. Xinyang, Jiujiang, Nanchang and Yiyang are medium-sized cities neighboring Hubei. In particular, Xinyang appears to be in a particular vulnerable situation because of the large number of people traveling from Wuhan and its insufficient healthcare resources.

## Discussion

The reported vulnerability analysis informs public health response to the 2019-nCoV epidemic in multiple ways. For the identified cities with high vulnerability, it is important to increase the reserve of healthcare resources or have contingent plans. In particular, some of these cities already contributed significant healthcare resources to Hubei, leaving themselves inadequately prepared for the localized outbreak. Our analysis revealed that several medium-sized cities, particularly those neighboring Hubei, are highly vulnerable due to the lack of healthcare resources.

## Data Availability

All data are available online (as indicated by the links in the manuscript).

---

**Figure 1.**
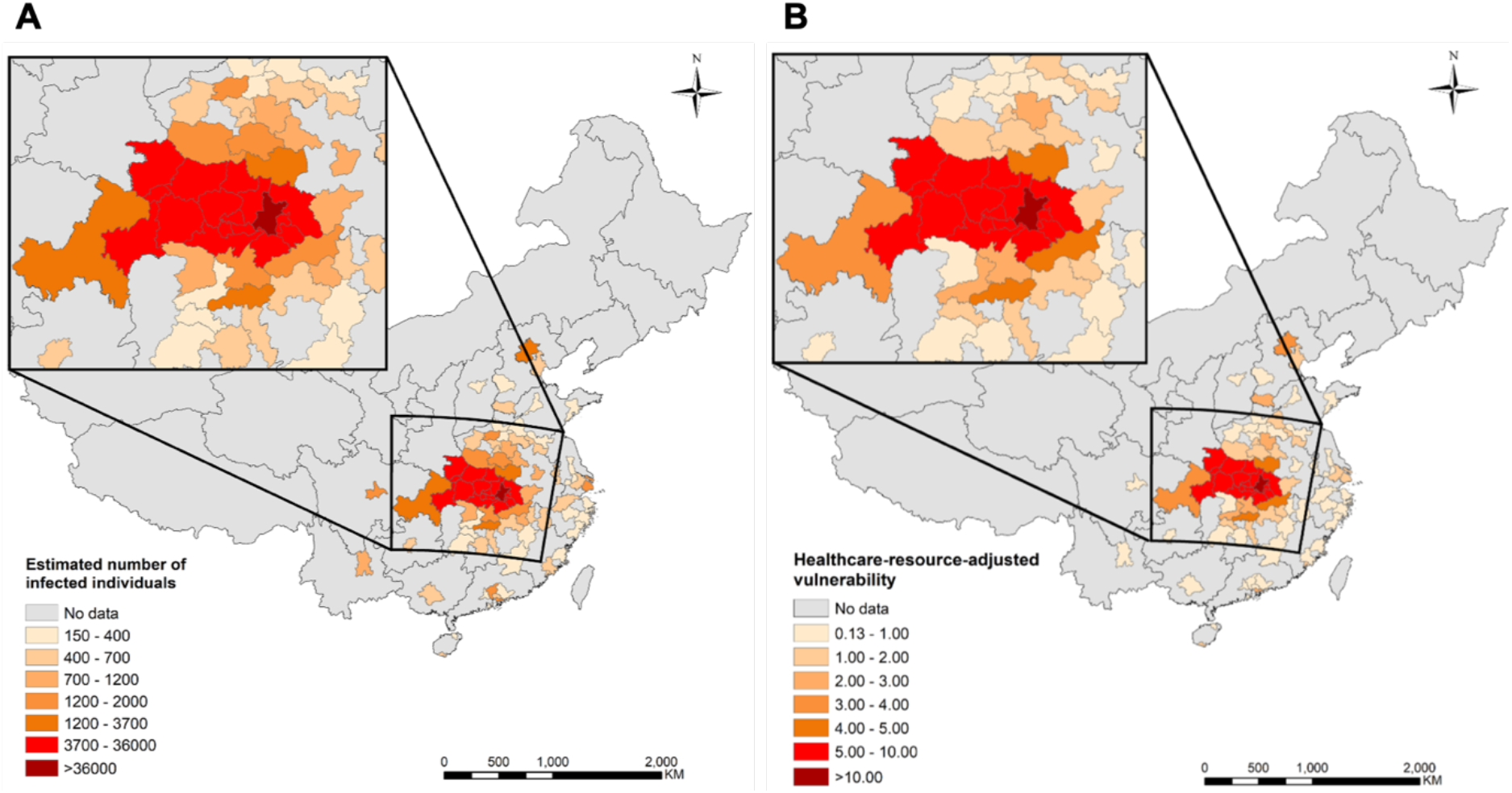
(A) The estimated number of infected individuals. (B) The healthcare-resource adjusted vulnerability to 2019-nCoV outbreak across mainland China. (C) The ten most vulnerable cities outside Hubei (January 29, 2020).

**Table 1.** The ten most vulnerable cities outside Hubei (January 29, 2020).

| City | Province/<br>Municipality | Total number of<br>reserved beds in<br>designated hospitals<br>$B_i$ | Estimated number<br>of 2019-nCoV<br>infected<br>individuals<br>$ID_i$ | Vulnerability<br>$V_i = ID_i / B_i$ |
| --- | --- | --- | --- | --- |
| Xinyang | Henan | 233 | 3230 | 13.84 |
| Changsha | Hunan | 655 | 4042 | 6.17 |
| Beijing | Beijing | 763 | 4357 | 5.71 |
| Jiujiang | Jiangxi | 317 | 1453 | 4.58 |
| Shenzhen | Guangdong | 500 | 2123 | 4.25 |
| Chongqing | Chongqing | 1074 | 3384 | 3.15 |
| Shanghai | Shanghai | 1060 | 3134 | 2.96 |
| Nanchang | Jiangxi | 634 | 1558 | 2.46 |
| Suzhou | Jiangsu | 384 | 931 | 2.43 |
| Yiyang | Hunan | 145 | 351 | 2.42 |

